# Associations between sustainable development goals accelerators and well-being, by household heads’ disability status among adolescents in Zambia – a cross-sectional study

**DOI:** 10.1101/2021.10.22.21264347

**Authors:** David Chipanta, Janne Estill, Heidi Stöckl, Lucas Hertzog, Elona Toska, Patrick Chanda, Jason Mwanza, Kelly Kaila, Chisangu Matome, Gelson Tembo, Olivia Keiser, Lucie Cluver

## Abstract

**Objectives:** We examined associations between accelerators (interventions impacting two or more SDG targets) and well-being indicators among adolescents in Zambia.

**Methods:** We randomly sampled 1,800 households receiving social cash transfers (SCT) in four districts, surveyed adults 16 years and older. Using multivariable logistic regressions, stratified by household heads disability status, we examined associations between accelerators (SCT, life-long learning (LLL), mobile phone access (MPA)) and seven well-being indicators among adolescents 16 to 24 years old. We predicted adolescents’ probabilities of reporting indicators using marginal effects models.

**Results:** We included 1,725 adolescents, 881(51.1%) girls. MPA was associated with no poverty (adjusted Odds Ratio [aOR] 2.08, p<0.001), informal cash transfers (aOR 1.82 p=0.004), seeking mental support (aOR 1.61, p=0.020); SCT with no health access restrictions related to disability (aOR 2.56, p=0.004), lesser odds seeking mental support (aOR 0.53, p=0.029); LLL with informal cash transfers (aOR 3.49, p<0.001), lower school enrolment (aOR 0.70, p=0.004). Adolescents living with disabled household heads reported worse poverty, good health, less suicidal ideation.

**Conclusions:** Accelerators - SCT, LLL, MPA - were associated with well-being indicators. Adolescents living with disabled household heads benefited less.

**Relevance to SDGs:** This paper shows that adopting accelerators can help achieve SDGs-aligned well-being indicators for adolescents living in poverty. However, accelerators may not offset disability-related inequalities. Adolescents living with disabled household heads may require more attention to achieve the SDGs.

**SDGs targets:** 1.2. no poverty; 1.3.1 social cash transfers, Informal cash transfers; 3. good Health; 3.4. no suicidal ideation; 3.4. seeking mental support; 4.1. school enrolment; 10. no health access restrictions related to disability.

## Introduction

Adolescents are a crucial population group to attain the Sustainable Development Goals (SDGs). Individuals aged 15 to 24 years comprise 15.5% (1.21 billion) of the global population, reaching 1.29 billion by 2030 [1]. Adolescence is a decisive period to intervene on multiple SDGs. The rapidly developing physical and mental growth, transition into adulthood taking place during adolescence have strong impact on health and well-being in adulthood [2,3]. In sub-Saharan Africa, where the growth of adolescents’ population is fast, the potential to improve their well-being is more constrained [1]. The region’s adolescents have high rates of mental health conditions, suicide, HIV, and other diseases [4]. A 10-year-old child is six times more likely to die by age 24 in sub-Saharan Africa than in North America or Europe. Globally, suicide is the second leading cause of death among adolescents aged 15 - 25 years [4]. Suicidal ideation, defined as a preoccupation with thoughts of killing oneself, and planning of suicide among adolescents aged 13–17 years in low-income and middle-income countries were the highest in Africa [5]. Not being in employment, education, or training (NEET) also negatively impacts adolescents’ well-being and successful transition into adulthood [6]. A quarter (25.9%) of adolescent girls and 15.8% of boys in sub-Saharan Africa were NEET in 2019 [7]. Of those employed, the majority (94.9 %) were in informal employment, living in extreme poverty, on less than US$1.90 a day [7]. Mobile phone use which is among interventions that could improve adolescents’ achievements of the SDGs is also limited in the region [8,9].

Urgent government coordinated actions are needed to accelerate the achievement of SDGs for the regions’ adolescents, particularly in the context of the Coronavirus Disease 2019 (COVID-19) pandemic [10]. The United Nations Development Programme (UNDP) defines accelerators as interventions or circumstances that positively impacts two or more SDGs targets [11]. Studies on accelerators have shown multiple, large, and favourable changes in adolescents’ mental health, the experience of transactional sex, violence, HIV prevention and treatment, and other SDG aligned outcomes from combining and re-arranging existing interventions. For example, social protection including cash transfers, education, safe schools, food security, parenting programmes, role of caregivers, and psychosocial support have been shown to be accelerators [12-14].

Disability is a serious threat to achieving the SDGs [15]. More than 1 billion people worldwide are estimated to be living with disabilities. The majority are left behind in several SDGs [15,16]. On the other hand, cash transfer programmes, in general not only include people with disabilities [15], but also often pair the programmes with training (life-long learning) to emphasize or explain programme objectives. The programmes also deliver cash and other services via mobile phones to individuals and households [17]. Social cash transfers (SCT), life-long learning (LLL) and mobile phone access (MPA) could potentially be accelerators and support adolescents in households headed by persons with disabilities. We, therefore, aimed to test whether SCT, LLL and MPA fulfil the definition of accelerator in this study, and how they interact with the household heads’ disability status in improving the SDGs aligned well-being indicators for adolescents.

## Methods

### Data sources and sample

We used the baseline data collected in August to September 2019 for the evaluation of the United Nations Partnership on the Rights of People with Disabilities (UNPRPD) project in Luapula province [18]. The UNPRPD started in January 2019 in two Luapula province districts in Zambia and will end in December 2021. It aims to increase HIV and sexual and reproductive health services among girls and women with disabilities receiving SCT in two districts. We also collected data from two districts in the same province not covered by the UNPRPD, but receiving SCT, to provide comparators in the evaluation.

Households are eligible to receive the SCT if government authorities identify them as extremely poor through measures of standards of living and satisfying one or more of the following criteria: women-headed; headed by a person aged 65 years or older, have a member with a disability; have adult members who are unable to work or support themselves economically and host orphans and vulnerable children, i.e., any child below 18 years who may be living with HIV, has lost one or both parents to HIV, or from any cause, or lives in a community affected by HIV [19]. Eligible households received ZMK90 (USD12) per month, and ZMK180 (USD24) if they included a person with a disability. The payments were disbursed every two months through a local pay point manager, the post office, or the recipients’ bank account [19].

### Sample size calculation

We calculated a minimum sample size of 1,800 households, from 90 community welfare action committees (CWACs) which are political units, and 20 households per CWAC. Our sample size calculation assumed a statistical significance (α) of 0.05, power of 80% and Intra-class Correlation Coefficients (ICCs) (p) of 0.01 to 0.08 and intervention effect (□) of at least 0.20 on HIV prevention services including condom use [20]. We sampled respondents in two stages. In stage one, we sampled CWACs using proportional probability sampling without replacement so CWACs with more households and typically with more services would be more likely to be selected. In stage two, from each CWAC, we sampled 25 households, instead of 20, to allow for non-response.

### Procedures

Trained fieldworkers first obtained and recorded consent from every respondent aged 16 years or older on the electronic tablets (thumbprints for oral, and signatures for written consents). They then administered a questionnaire in the area’s local language on the electronic tablets installed with Open Data Kit software to the household head and all household members aged 16 years or older who consented. The survey contains questions on socio-demographic characteristics, self-rated poverty, health and well-being, mental health, school enrolment, disability status, proximity to health facilities, health access restrictions related to disabilities, health services, receipt of SCT offered by the government, non-governmental organizations and individuals, training, and MPA. We derived the questions from piloted and validated tools, including the UNICEF Innocenti tools and Demographic and Health Survey. We translated the questions from English into the local language. We trained the fieldworkers using role plays to ensure understanding and standardized administration of the questionnaire. We stored and electronically transferred the data to a secure server. We analyzed responses only from respondents aged 16 to 24 years.

The study protocol was reviewed by the University of Zambia Humanities and Social Sciences Research Ethics Committee (IRB Approval No. 2019-April-001) and the ethics committee in the Canton of Geneva (no 2019-00500).

### Measures and variables

We identified and three potential accelerators: 1) SCT, 2) LLL, and 3) MPA, and seven SDGs aligned indicator target outcomes in the data based on our review of the literature. We defined the accelerators as follows: SCT provided by the government with the question: “During the past 12 months, has the respondent or any household member received money or goods, including food, clothing, livestock, or medicines from any of the following government programmes, social cash transfers and other government transfers?” (Other government transfers combined respondents or their household’s receipt of school uniforms, scholarships, food security pack, school feeding, and farm input subsidy), coded no, yes; LLL, combined participation in government offered training on HIV, disability, gender-based violence, human rights, sexual and reproductive rights, job skills, social protection and economic empowerment derived from the question: *“*During the past 12 months, have you or any of the household members received any training provided by the government on general health, food and nutrition, sexual and reproductive rights, HIV, human rights, and gender-based violence, social protection, job skills, and economic empowerment?” coded yes if the participant responded to have participated in any of the training, otherwise no; MPA with the question *“*What phone number is used at this house?” (Response options were no phone, phone number)” coded no for no phone, yes for phone number.

We defined the SDG aligned indicator outcomes as follows: No poverty with the question: *“*Do you consider your household to be nonpoor, moderately poor, or very poor?” coded very poor, moderately poor; Informal cash transfers with the question: “During the past 12 months, has the respondent or any household members received money or goods, including food, clothing, livestock, or medicines from individuals who are not part of the family or non-governmental organizations?” coded no, yes; Good health with the question: *“*Have you been sick or injured in the last two weeks?” coded physically sick, not sick; No suicidal ideation with the question: “Did you have thoughts of hurting or killing yourself? coded *yes, no*; Seeking mental support with the question “What health facility or other institutions or persons did you see for any of the identified mental health issues?” coded no did not see; yes saw. Seeking mental support proxied having mental health problems and seeking help to resolve them. School enrolment combined the responses from the question: *“*Are you currently attending school? (Check relevant choice) nursery/pre-school, other grades full-time, other grades part-time, community school, full-time, correspondence, adult literacy class, tertiary school” coded no, yes. The proportion of adolescents currently in school versus those not in school for 20 to 24-year-olds, coded no, yes; No health access restrictions related to a disability with the question: *“*Are you limited in accessing health services because of your impairment?” coded limited, not limited.

We controlled for age (16 to 19, 20 to 24 years), biological sex (male, female), disability status (not disabled, disabled), proximity to health facility (<7 or ≥ 7 kilometres), and district (Kawambwa, Nchelenge, Mansa, and Samfya). We assessed household heads’ disability with questions from the Washington Group Short Questions (WGSQ) on disabilities. The WGSQ asks respondents if they have difficulties with seeing, hearing, walking, cognition, self-care, or communicating. For each disability type, the answer options are “no”; “yes – a little”; “yes – a lot”; “cannot at all.” We defined a respondent as disabled in the disability type they answered: “yes – a lot” or “cannot at all” and grouped the disability variables into a composite variable reflecting if the respondent had any of the six types of disability [21].

### Analysis

We conducted analyses in three steps. First, we explored the socio-demographic characteristics, hypothesized accelerators and SDGs aligned outcomes by household head’s disability status. Second, we tested for associations between each SDG-aligned outcome and hypothesized accelerators simultaneously using the Fishers exact test and reported crude proportions, 95% confidence intervals (CI) and p-values. We adjusted for covariates in multivariable logistic regressions and corrected for multiple hypothesis testing using the Benjamin, Yekutieli, Krieger (BYK) False Discovery Rate Sharpened Qs [22]. We interpreted the FDR adjusted p-value as a p-value of 0.05, resulting in 5% of significant tests being false positives. FDR adjusted p-values result in fewer false positives than non-FDR adjusted p-values. Third, we predicted adolescents’ probabilities of experiencing each outcome from no accelerators to cumulative accelerators combinations by household heads’ disability status using marginal effects models with the Stata margins command keep other covariates at their mean values. We reported the changes in probabilities for each indicator.

As a sensitivity analysis, we calculated adjusted probabilities of experiencing each outcome from multiple-outcome probit models that correlated the error terms of three potential accelerators in each model, using the mvprobit command in Stata 14.1 set at 50 random draws. Each regression regressed one of the seven SDG aligned outcomes for adolescents on the three accelerators controlling for sociodemographic factors. We clustered analyses at the CWAC level and used Stata version 14.1 for analysis.

## Results

The sample comprised 1,725 adolescents from 1,545 households in 90 CWACs. Overall, 881 (51.1%) were girls and 844 (48.9%) boys. The median age in years was 19 (interquartile range 17 to 21). Eight per cent (145) of the adolescents lived with household heads with disabilities. Half (75, 51.7%) of household heads with disabilities reported at least “a lot” of difficulties in remembering, 43 (29.6%) in seeing and 14 (9.6%) in self-care. Many socio-demographic characteristics and SDG aligned targets indicators differed significantly between adolescents living with household heads with and without disability; the three hypothesized accelerators did not differ.

The three hypothesized accelerators - SCT, LLL and MPA – were significantly associated with no poverty, informal cash transfers, good health, no suicidal ideation, school enrolment and no health access restrictions related to disabilities when we did not control for socio-demographic covariates. However, SCT was associated with lower levels of seeking mental support among adolescents (Table 2A). Adolescents with MPA, reported higher levels of no poverty (39% versus 23.9%, p <0.001), accessing informal cash transfers (26.6% versus 16%, p<0.001), good health (34.5% versus 30.2%, p=0.042), seeking mental support (38.4% versus 26.9%; p<0.000) and school enrolment (48.8% versus 39.7%, p <0.001) than those without MPA.

**Table 1:**
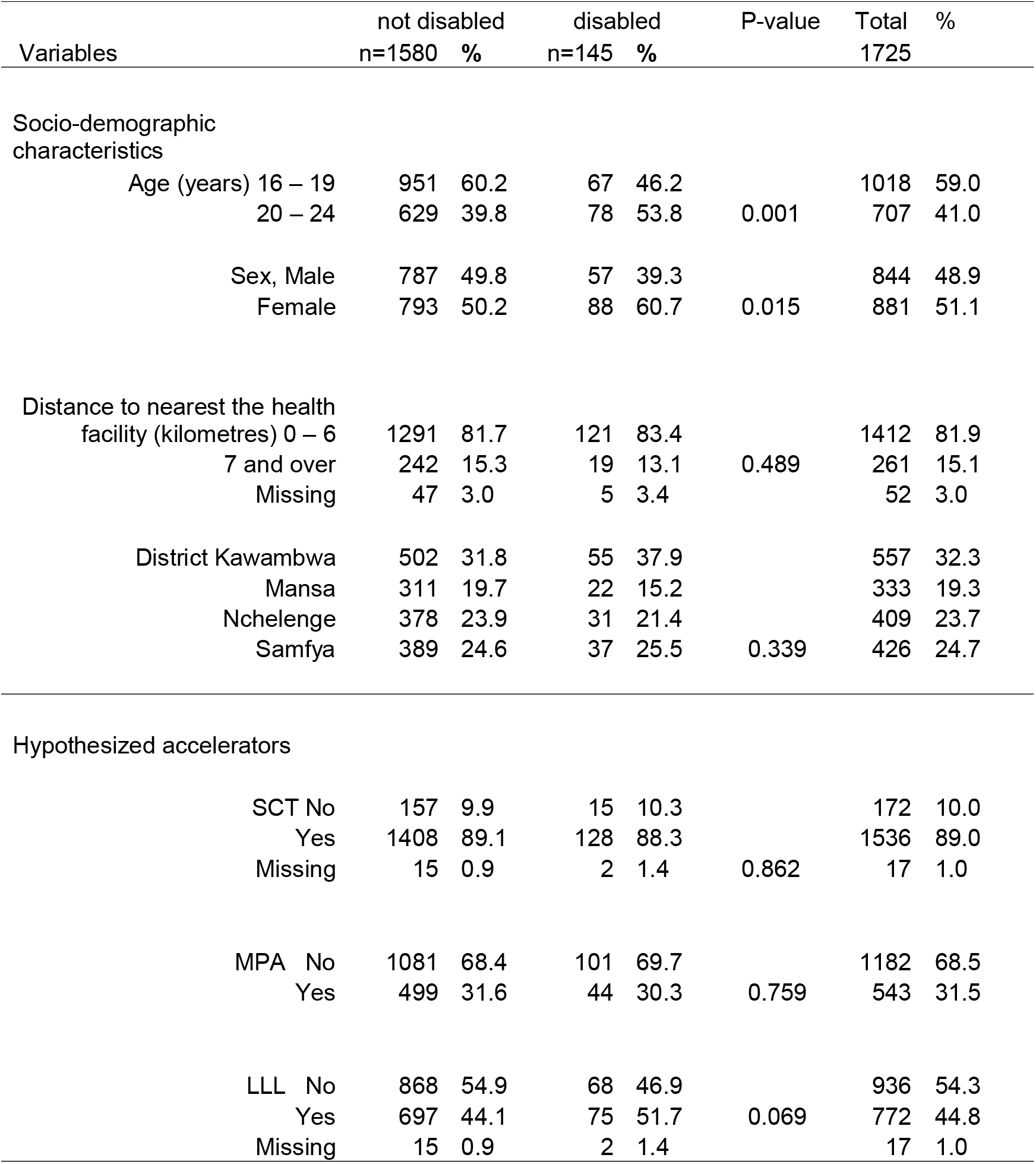
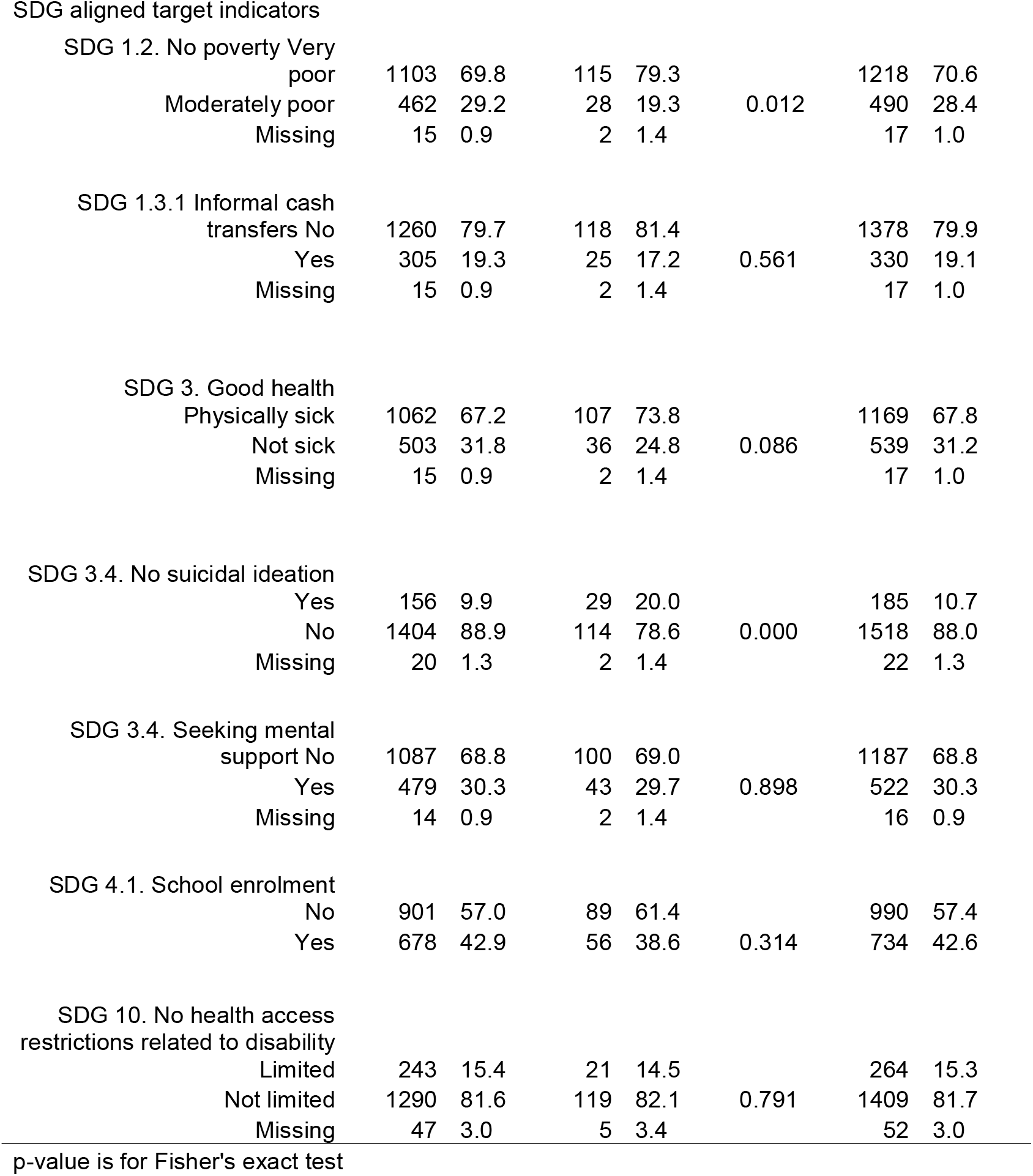
Social demographic characteristics, hypothesized accelerators and SDG-aligned targets by disability status of the household head

**Table 2A:** Crude analysis of associations between hypothesized accelerators and SDG aligned targets

| SGD-aligned targets | *Hypothesized accelerators, absolute values [proportions], p-value* |  |  |  |  |  |
| --- | --- | --- | --- | --- | --- | --- |
|  | SCT |  | LLL |  | MPA |  |
|  | Yes | No | Yes | No | Yes | No |
| 1.2. No poverty | 442 [28.8%], | 46 [26.7%]; 0.318 | 207[26.8%], | 281 [30.1%]; 0.074 | 212[39.0%], | 278 [23.9%]; <0.001 |
| 1.3.1 Informal cash transfers | 323 [21.0%], | 7[4.7%]; <0.001 | 228[29.5%], | 102 [10.9%]; <0.001 | 144[26.6%], | 186 [16.0%]; <0.001 |
| 3. Good health | 489 [31.8%], | 50 [29.7%]; 0.258 | 253[32.8%], | 285 [30.5%]; 0.168 | 187[34.5%], | 352 [30.2%]; 0.042 |
| 3.4. No suicidal ideation | 1381[90.2%], | 137 [79.7%]; <0.001 | 674[87.6%], | 843 [90.4%]; 0.044 | 475[87.6%], | 1043 [89.8%]; 0.102 |
| 3.4 Seeking mental support | 447 [29.1%], | 75 [43.6%]; <0.001 | 278[36.0%], | 244 [26.1%]; <0.001 | 208[38.4%], | 314 [26.9%]; <0.001 |
| 4.1. School enrolment | 662 [43.1%], | 65 [37.8%]; 0.103 | 302[39.1%], | 424 [45.4%]; 0.005 | 265[48.8%], | 469 [39.7%]; <0.001 |
| 10. No health restrictions related to disability | 1293[85.8%], | 116[69.9%]; <0.001 | 613[82.0%], | 795 [86.0%]; 0.014 | 453[83.9%], | 956 [84.4%]; 0.425 |

Fisher's exact test.

After adjusting for age, gender, household heads’ disability status, distance from the nearest health facility and district, all hypothesized accelerators remained associated with two or more SDG-aligned outcomes (Table 2B). Good health and no suicidal ideation were no longer associated with any hypothesized accelerator. Having access to a mobile phone was associated with higher odds of no poverty, accessing informal cash transfers, seeking mental support and school enrolment. SCT were associated with higher odds of informal cash transfers, no health access restrictions related to disability but lower odds of seeking mental support. LLL was associated with increased odds of accessing informal cash transfers but lower odds of school enrolment.

**Table 2B:**
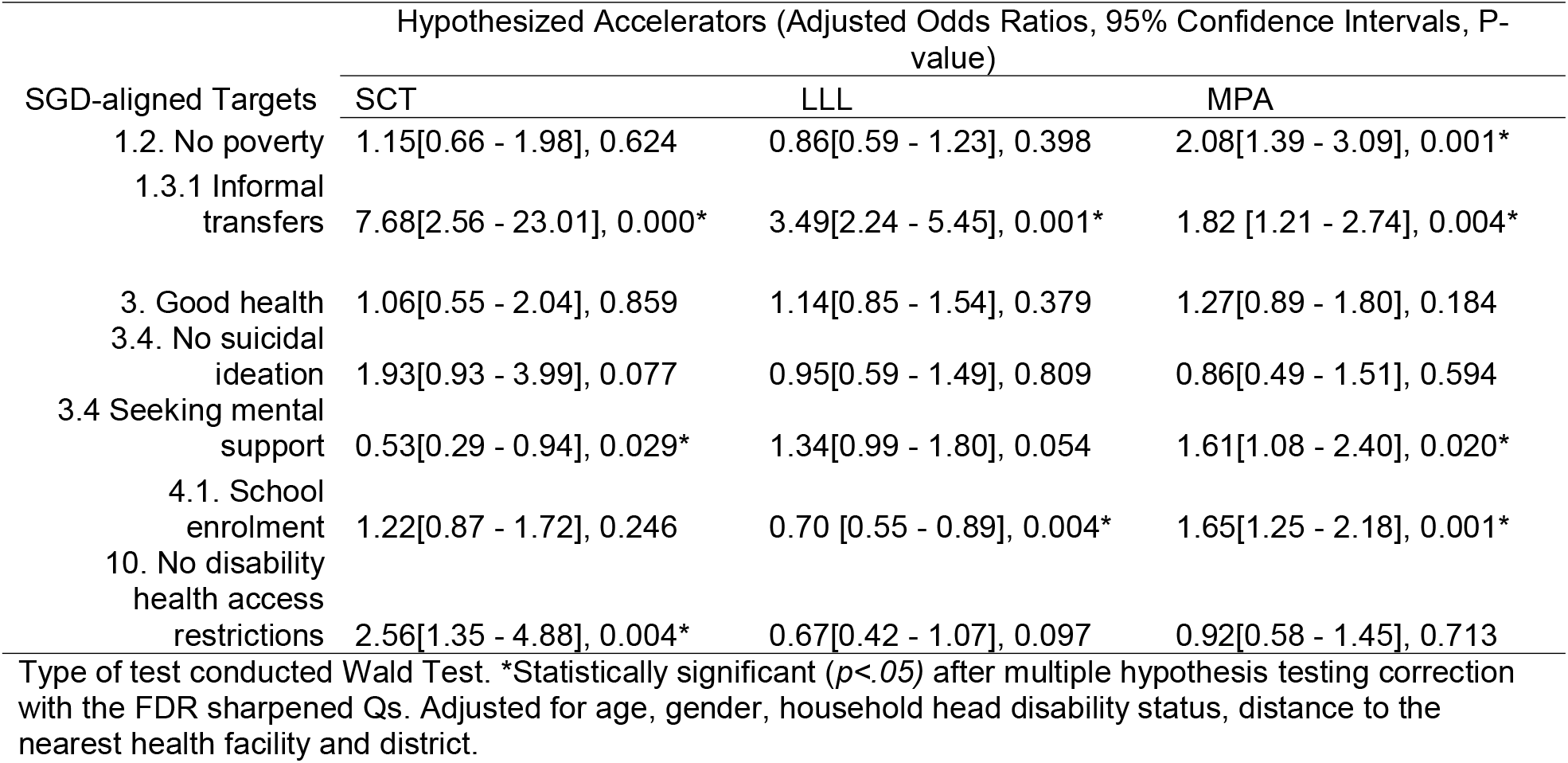
Associations between hypothesized accelerators and SDG aligned targets indicators adjusted for social demographic characteristics

| SGD-aligned Targets | Hypothesized Accelerators (Adjusted Odds Ratios, 95% Confidence Intervals, P-value) |  |  |
| --- | --- | --- | --- |
|  | SCT | LLL | MPA |
| 1.2. No poverty | 1.15[0.66 - 1.98], 0.624 | 0.86[0.59 - 1.23], 0.398 | 2.08[1.39 - 3.09], 0.001* |
| 1.3.1 Informal transfers | 7.68[2.56 - 23.01], 0.000* | 3.49[2.24 - 5.45], 0.001* | 1.82 [1.21 - 2.74], 0.004* |
| 3. Good health | 1.06[0.55 - 2.04], 0.859 | 1.14[0.85 - 1.54], 0.379 | 1.27[0.89 - 1.80], 0.184 |
| 3.4. No suicidal ideation | 1.93[0.93 - 3.99], 0.077 | 0.95[0.59 - 1.49], 0.809 | 0.86[0.49 - 1.51], 0.594 |
| 3.4 Seeking mental support | 0.53[0.29 - 0.94], 0.029* | 1.34[0.99 - 1.80], 0.054 | 1.61[1.08 - 2.40], 0.020* |
| 4.1. School enrolment | 1.22[0.87 - 1.72], 0.246 | 0.70 [0.55 - 0.89], 0.004* | 1.65[1.25 - 2.18], 0.001* |
| 10. No disability health access restrictions | 2.56[1.35 - 4.88], 0.004* | 0.67[0.42 - 1.07], 0.097 | 0.92[0.58 - 1.45], 0.713 |

Type of test conducted Wald Test. \*Statistically significant ( $p < .05$ ) after multiple hypothesis testing correction with the FDR sharpened Qs. Adjusted for age, gender, household head disability status, distance to the nearest health facility and district.

Figure 1 shows the changes in probabilities of experiencing each of the seven SDG-aligned outcomes from potential accelerators compared to no accelerators: 1) SCT alone, 2) SCT plus LLL, 3) SCT plus MPA, 4) SCT, plus LLL and MPA. Results are stratified by disability status of the household head. Potential accelerators were associated with an absolute increase of at least 0.02 in the probability of SDG aligned targets indicators. However, the probability of seeking mental support was decreased by SCT alone, SCT plus LLL, and SCT plus MPA. The probability of school enrolment was also decreased by SCT plus LLL.

**Figure 1:**
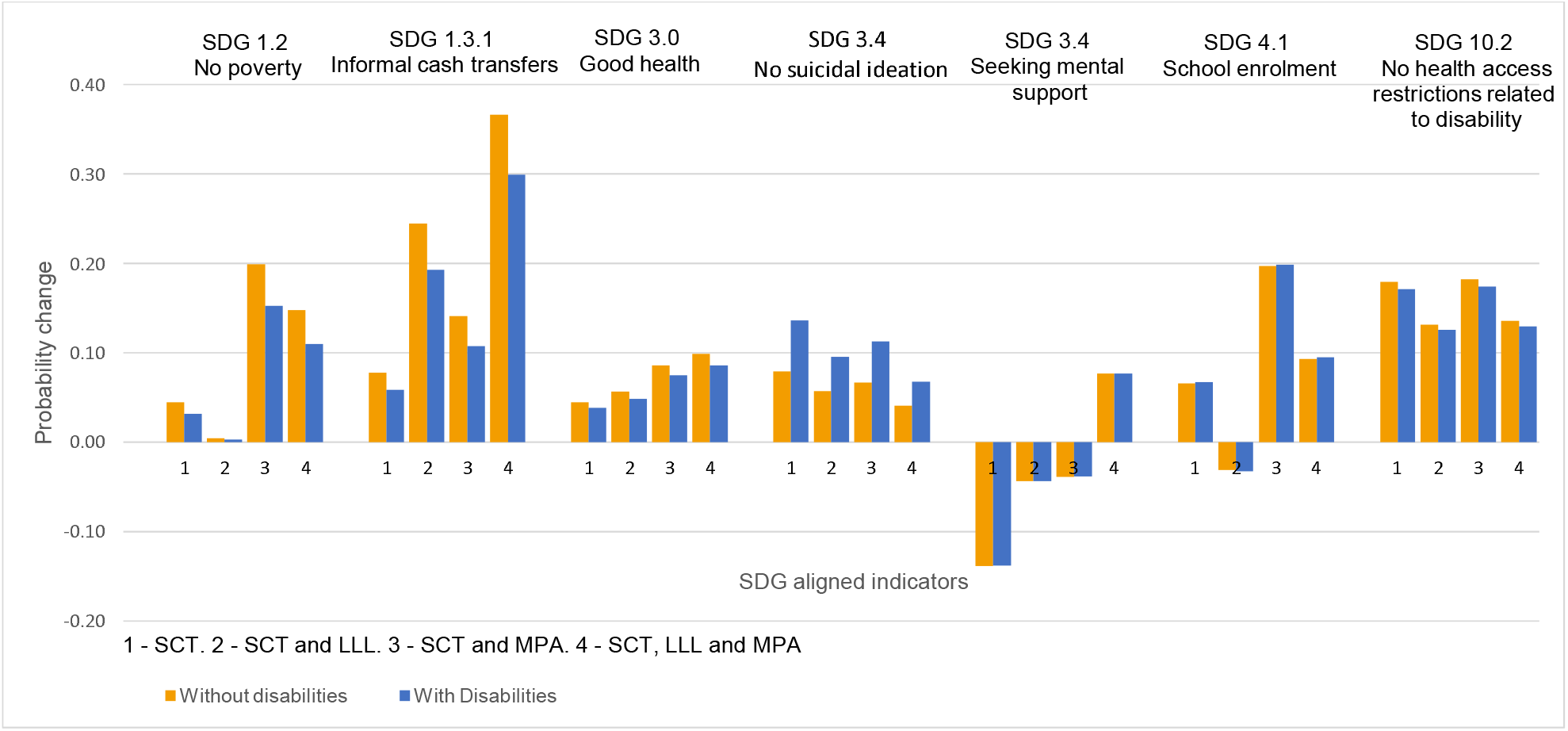
Levels of probability change in SDG-aligned targets indicators outcomes from 1) SCT alone, 2) SCT plus LLL 3) SCT plus MPA, and 4) SCT plus LLL and MPA, stratified by household heads’ disability status – without (blue bars) and with disabilities (Orange bars).

Adolescents with household heads with disabilities had lower probabilities of reporting no poverty, accessing informal cash transfers, good health and no suicidal ideation from no potential accelerators than their counterparts without household heads with disabilities. They further reported lower probability changes from potential accelerators in no poverty, accessing informal cash transfers, good health, and no health access restrictions related to disability. The probability increase in no suicidal ideation from potential accelerators was higher among adolescents living with household heads with disabilities. Changes in seeking mental support and school enrolment did not differ by the disability status of the household head. The greatest probability changes from receiving no potential accelerators to receiving potential accelerators were in accessing informal cash transfers (Figure 1).

Synergies – combinations – of potential accelerators were associated with changes in the probabilities of experiencing levels of SDG-aligned targets indicators outcomes for adolescents living with household heads with and without disabilities. A combination of all potential accelerators - SCT, LLL and MPA – was associated with a 0.15 and 0.11 probability increase in levels of no poverty for adolescents living with household heads without and with disabilities; 0.37 and 0.30 probability increase in levels of accessing informal cash transfers and 0.14 and 0.13 of experiencing no health access restrictions related to a disability, respectively (Figure 2).

**Figure 2:**
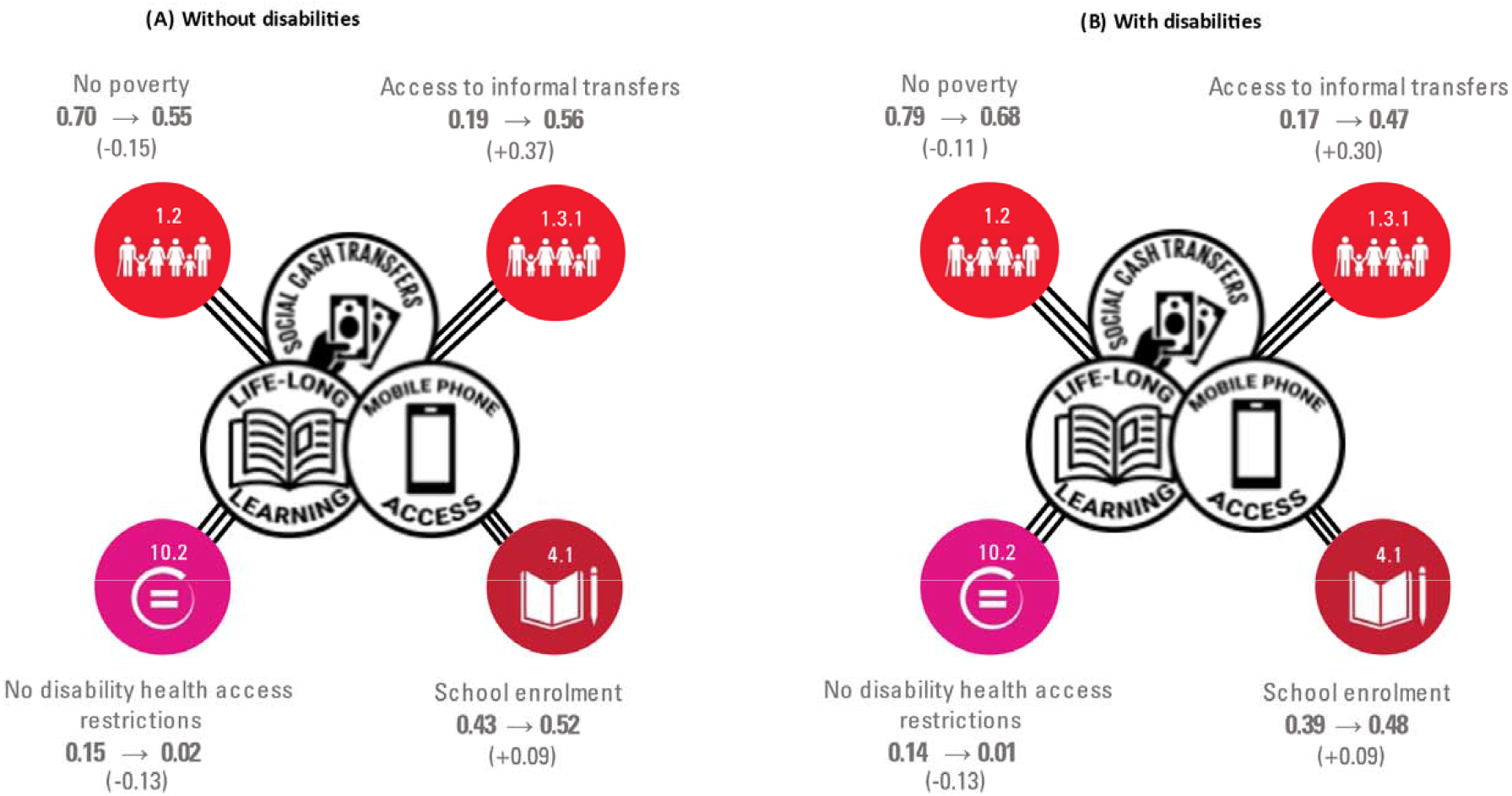
Changes in probability levels of SDG-aligned outcomes for adolescents living with household heads without (A) and with disabilities (B) from synergies of interventions.

The sensitivity analysis results between the models we used and the models that account for correlations between the error terms of the potential accelerators were equivalent. However, the p-values were lower in the outcome models accounting for the correlation between potential accelerators (Supplementary Table 1).

## Discussion

This study examined associations between potential accelerators - SCT, LLL, and MPA - and SDG aligned well-being indicator targets - 1.2 no poverty; 1.3.1 SCT, Informal cash transfers; 3.0 good Health; 3.4 no suicidal ideation, seeking mental support; 4.1 school enrolment; and 10.0 no health access restrictions related to disability - among adolescents. We found high potential for improving vulnerable adolescents’ SDG-aligned well-being by combining SCT, LLL and MPA interventions. Our findings fit within an emerging body of evidence confirming that SCT, LLL and MPA are accelerators for adolescents [8,10,13]. It further found that adolescents benefited unequally depending on their household heads’ disability status and that combining existing interventions may not overcome inequalities arising out of the disability of the household head.

Several studies conducted in Zambia and elsewhere confirm our results that SCT were associated with multiple SDG aligned target outcomes such as higher levels of informal cash transfers, no health access restrictions related to disability, and lower levels of seeking mental support. Studies conducted in Zambia show that SCT reduced relative poverty, increased women’s satisfaction regarding their children’s well-being, and schooling among school-going adolescents [23,24]. SCT also increased material well-being (children’s material needs met), food insecurity, and asset ownership [25]. In sub-Sahara Africa and elsewhere, SCT have been shown to increase psychological well-being, and decrease relative and absolute poverty [26,27]. In our study, receiving SCT alone was associated with a substantial decrease in seeking mental support. Combining SCT with LLL or MPA was associated with more reductions in seeking mental support. This result suggests that households’ lack of money, LLL opportunities, and MPA may have necessitated respondents to seek mental support. Providing SCT, LLL and MPA interventions may be vital for addressing the mental support needs of adolescents living in poverty.

However, in our study SCT were not associated with good health; neither were MPA and LLL. This result fits within a body of evidence showing that cash transfers have positive, complex, and mixed effects on health. A review of 56 studies from low and middle-income countries found that cash transfers increased dietary diversity, access and utilization of health services but had little impact on children’s anthropometric measures [26]. In high-income countries, self-rated health, chronic health conditions, and mortality for cash transfer recipients were worse than among non-recipients. On the contrary, in the United States, cash transfers were associated with improved self-rated health [28]. One reason why SCT, MPA, and LLL in our study were not associated with good health could be that physical illnesses among our sample was widespread. Two-thirds (67.8%, n=1169) of adolescents reported physical illnesses. Another is that malaria is endemic in the study area [29]. SCT, MPA and LLL alone might have been insufficient to resolve these illnesses. Innovative prevention and treatment of malaria, and other illnesses, combined with SCT, MPA, and LLL, should be implemented.

Contrary to views that cash transfers and other public transfers reduce informal transfers [30], our study found the opposite result; SCT were associated with increased receipt of informal cash transfers. This result is supported by evidence [31,32]. One explanation for our study’s finding is that the process of receiving SCT may have identified households who were in need of financial and material support, linked them to support, strengthened trust of each other, increased social inclusion and solidarity [31-33]. Another explanation is that our study did not include pensions and social security transfers analyzed in the study that found contradictory findings to our study results [30]. Pensions and social security transfers, derived from mandatory savings employees make during employment, tend to be larger than SCT. In addition, pensions and social security transfers recipients may be wealthier, making them less likely to be perceived as in need of informal cash transfers [31].

The positive associations found in our study between MPA and no poverty, informal cash transfers, and school enrolment are also supported by evidence [8,34]. Access to mobile phones can promote adolescents’ wellbeing, expand their social networks and personal growth opportunities [8]. Social protection and cash transfers are being delivered via mobile phones, alongside electronic vouchers, bank accounts and other payment systems to adults in households [35]. Mobile phones use also enables access to vital services [8,34]. Informal financial transfers make the bulk of financial transactions transferred via mobile phones in sub-Sahara Africa [36]. The negative association found in our study between MPA and seeking mental support suggests that lack of mobile phone access may be mentally distressing for adolescents. One main reason is that they may miss out on informal cash transfers and other services to improve their well-being [8,34,36]. Providing mobile phones to households who do not have them, is being done and can help improve adolescents access to social protection, cash transfers and mental support [35].

In our study, LLL’s associations - increase in informal cash transfers, and reductions in odds of seeking mental support and school enrollment – are limited than those of SCT and MPA, but no less critical. LLL re-enforces and complements the objectives of social protection and cash transfer programmes. LLL may bring participants together, potentially increasing their social networks - addressing the needs to seek mental support - and informal cash transfers. LLL is unlikely to have pulled adolescents out of school. Two-thirds of adolescents were already not attending school. LLL and the skills it provides can be beneficial to these adolescents [37].

Adolescents did not evenly benefit from SCT alone, with LLL, MPA or LLL and MPA, although they benefited from these accelerators. Adolescents living with household heads with disabilities reported lower benefits from these accelerators in no poverty, informal cash transfers, good health, and no disability health access restrictions than those living with household heads without disabilities. Previous studies, including a systematic review, support this finding showing that living with a household member with a disability had high cost and poverty implications for the household [38,39]. These studies concluded that households must spend as much as 26% more resources to obtain an equivalent standard of living compared to those without disabilities [38,39]. Adolescents in our sample living with household heads with disabilities reported themselves poorer, may have had much more diminished resources and saddled with caregiving responsibilities adversely affecting their well-being than their peers living with household heads without disabilities. However, adolescents living with household heads with disabilities reported greater benefits from accelerators in no suicidal ideation. Their probabilities of reporting no suicidal ideation from no accelerators were lower compared to their peers without household heads with disabilities. This result suggests that household heads’ disability status may have mitigated suicidal ideation among adolescents. Such adolescents might have benefited from parental supervision during caregiving which is known to be protective against suicidal behaviour [40]. However, this study did not look at the role of the household head’s disability status on adolescents’ suicidal ideation. Overall, accelerators appear to impact adolescents’ well-being. However, adolescents living with household heads with disabilities were doing worse than their peers living with household heads without disabilities before and after the accelerators. New interventions focused at households may be required. These could include attention to adolescents and parents’ relationships, increased psychosocial, mental and financial support to offset adolescents’ household’s disabilities-related inequalities [38].

## Limitations

It is important to note that association does not imply causal relationships. In this study we could not attribute causation due to the study’s cross-sectional nature, and neither could we generalize the results outside the study area and population group. However, many countries in sub-Saharan Africa are implementing similar programmes and could find our results useful in their contexts. We performed a complete sample analysis due to the low prevalence of disability in our sample, which might have missed nuanced differences experienced by adolescent girls compared to boys. The prevalence of disability in our sample was low limiting our ability for further analysis by type of disability. We did not have variables on occupational type of the household head, and others which could affect household dynamics including adolescents’ well-being. We did not input the missing data because it was less than 5%. However, we show that adopting accelerators can help achieve SDGs-aligned well-being indicators for adolescents living in poverty.

## Conclusion

Our study found multiple and substantial benefits from accelerators – SCT, LLL and MPA – delivered individually and in combinations, on SDG aligned well-being among adolescents living in poverty. Adolescents living with household heads with disabilities benefited less. New interventions maybe necessary to correct disability-related inequalities between households. More research is needed to understand combinations of interventions that improve the well-being of adolescents living with household members who are disabled.

## Data Availability

The data set is not publicly available because it contains unique identifiers and respondents did not agree to have the details public, although they consented to the study.

---

**Supplementary Table 1:** Associations between hypothesized accelerators and SDG aligned targets using probit models

| SGD-aligned targets | Hypothesized accelerators, Coefficient [95% CI], P-value |  |  |
| --- | --- | --- | --- |
|  | SCT | LLL | MPA |
| 1.2. No poverty | 0.15 [-0.08 - 0.37], 0.197 | -0.14 [-0.27 - -0.01], 0.043 | 0.44 [0.30 - 0.57], <0.001 |
| 1.3.1 Informal cash transfers | 0.80 [0.44 - 1.17], <0.001 | 0.66 [0.51 - 0.81], <0.001 | 0.35 [0.20 - 0.50], <0.001 |
| 3. Good health | 0.14 [-0.08 - 0.36], 0.215 | 0.04 [-0.09 - 0.17], 0.549 | 0.13 [-0.01 - 0.26], 0.064 |
| 3.4. No suicidal ideation | 0.44 [0.20 - 0.69], <0.001 | -0.17 [-0.33 - -0.01], 0.048 | -0.08 [-0.26 - 0.09], 0.342 |
| 3.4 Seeking mental support | -0.44 [-0.65 - -0.23], <0.001 | 0.29 [0.16 - 0.42], <0.001 | 0.31 [0.18 - 0.45], <0.001 |
| 4.1. School enrolment | 0.17 [-0.04 - 0.40], 0.117 | -0.23 [-0.36 - -0.1], <0.001 | 0.31 [0.17 - 0.45], <0.001 |
| 10. No health restrictions related to disability | 0.65 [0.42 - 0.88], <0.001 | -0.22 [-0.37 - -0.08], 0.003 | -0.02 [-0.18 - 0.13], 0.776 |

Probit models accounting for correlation between error terms. Adjusted for age, gender, household head disability status, distance to the nearest health facility and district

